# Quantifying political influence on COVID-19 fatality in Brazil

**DOI:** 10.1101/2022.02.09.22270714

**Authors:** Leandro de Almeida, Pedro V. Carelli, Nara G. Cavalcanti, José-Dias do Nascimento, Daniel Felinto

## Abstract

The COVID-19 pandemic was severely aggravated in Brazil due to its politicization by the country’s central government. However, the impact of diffuse political forces on the fatality of an epidemic is commonly hard to quantify. Here we introduce a method to measure this effect in the Brazilian case, based on the inhomogeneous distribution throughout the national territory of political support to the central government. The correlation between fatality rate and political support grows as the government’s misinformation campaign is developed, leading to the dominance of such political factor for the pandemic impact in Brazil in 2021. Once this dominance is established, this correlation allows for an estimation of the total number of deaths due to political influence as 350 *±* 70 thousands up to the end of 2021.

## Introduction

The coronavirus disease 2019 (COVID-19) pandemic affected profoundly all regions in the globe. In Latin America, Brazil was hit in a particularly hard way [1], with strong criticism directed to the actions of its central government [2, 3, 4]. The effectiveness of individual measures by different governments to curb the pandemics is a current matter of debate and investigation [5, 6, 7, 10]. However, in some countries, such as Brazil and The United States [8, 9], broad political forces have opposed measures believed to be among the most effective, while promoting ineffective treatments [11]. The impact of diffuse political views on the death toll of the pandemic is typically difficult to quantify. However, due to the exacerbated role of this factor in Brazil, we show here that it is possible to estimate this number for Brazil with relatively low uncertainty, resulting in an excess of 350 ± 70 thousand deaths by the mid of November 2021, or about (57 ± 11)% of the total number of deaths. The key parameter allowing this estimation is the inhomogeneity of political support for the central government throughout the national territory, from which we extrapolate to obtain the number of deaths not influenced by this factor. Our analysis also reveals the temporal dynamics of such political risk aspects in Brazil, showing its increase during 2020 up to dominance in 2021.

Brazil is a country of continental dimensions with a population of more than 200 million people. It is based on a federation of 26 States plus a Federal District (see Fig. 1). These 27 administrative units independently elect each of their Governors, and the whole country elects a President to run its central government. The elections are direct and mandatory for all citizens aged 18 to 70. The last general election for Governors and President in Brazil occurred at the end of 2018, with Jair Messias Bolsonaro winning the presidency in the last round of votes on October 28th. The pandemic started at the beginning of the second year of their four-year mandates. As a country of great social inequality, the health assistance in Brazil is divided into two subgroups: the smaller private care (based on insurance and a variety of private health providers) and the National Unified Health System (SUS, from *Sistema Uénico de Sauéde*) that provides universal assistance. Although representing a significant improvement for the general Brazilian population, the SUS is chronically affected by structural problems, including gaps in organization and governance, low public funding, and suboptimal resource allocation [12]. Even though a great gap exists between the richest and poorest areas of the country, especially concerning access to more complex assistance in rural and remote areas, the SUS is present in the whole country. This health care system is administered by the three instances of executive power: Federal Government (national), States, and Municipalities. Through the Health Ministry, the Federal instance regulates and funds the SUS and also coordinates national health programs, such as vaccination. The States and Municipalities are the executors of the main aspects of the health care system. They are responsible for all levels of direct health assistance, from primary to tertiary. In order to understand the fatality dynamics of COVID-19 in Brazil, it is important to have in mind then that the States in Brazil, through their local health authorities, were responsible for determining and enforcing the most critical measures to curb the pandemics, like social distance and mask mandates, and generally followed the recommendations of the World Health Organization. These measures suffered direct opposition by the President, but were generally upheld by the Brazilian Supreme Court [2, 13].

**Fig 1.**
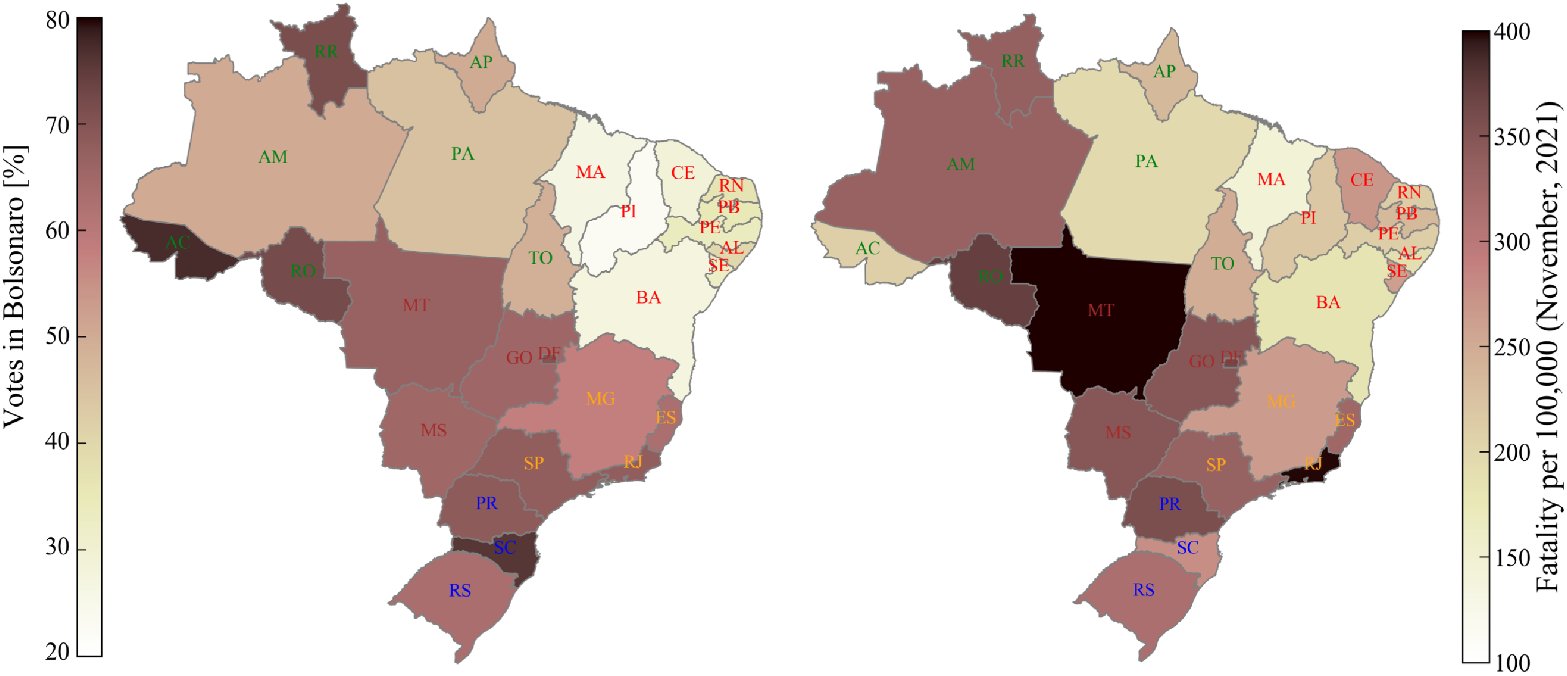
Geographical distributions of votes in Bolsonaro and of fatality by COVID-19 by mid-November 2021. Left map: colormap of votes in Bolsonaro per State in Brazil. States are labeled by their two-letter abbreviation. The label colors indicate the different regions of Brazil: Northeast (red), North (green), Central-West (Brown), Southeast (orange), South (blue), and Federal District (magenta). Right map: colormap of fatality per 100,000 as in November 17th, 2021.

The political influence we are discussing here, thus, is not concerned directly with the enforcement of particular measures to contain the pandemics, as investigated in previous works [5, 6, 7], but with the broad political action of the Presidency in Brazil to curb the efforts of the States to control the pandemics [2]. This political action was spread in multiple arms: systematically downplaying the risks of the pandemics, opposing national measures of social distancing and mask mandates, promoting ineffective treatments, delaying vaccination, and finally misinforming the population on the importance of preventive measures and on the risks and benefits of vaccination [4, 14]. Many measures against the spread of the COVID-19 virus depend on the compliance of the general population for their success, and this compliance can be directly affected by individual political views [15, 16]. The overall action of the Presidency in Brazil influenced a significant portion of the population to mishandle many of the measures to control the pandemics.

In the sea of factors influencing the dynamics of the pandemics, in general, it is challenging to estimate the effects of the actions of a political group and its leader on the behavior of the population, and in particular their effects on the pandemics numbers, such as deaths. However, in the case of Brazil, with the accumulation of time and consolidation of the misinformation campaign, a high correlation between deaths and voting rates in the 2018 election has emerged, as shown in Fig. 1. This correlation indicates a disparity throughout the country in the COVID-19 fatality rates depending strongly on the level of support for the Brazilian President in its various regions. As a result, this inhomogeneity in the States distribution of death rates and political support could be used to infer what the death rates would be without such political factors. From these last quantities, direct estimations can be derived for the impact of the political factors on the overall pandemic fatality in Brazil.

## 1 Methods

The temporal dynamics of the COVID-19 fatality rate followed very distinct patterns in the different States of Brazil. However, the general trend of political correlation started to form along the year 2020. This trend is shown in Fig. 2, through the depiction of the distribution of fatality rates among all Brazilian States in a temporal sequence since the beginning of the pandemic, organized as a function of the voting rate in Bolsonaro. The different States are represented by their two-letter abbreviations, as with SP for S∼ao Paulo, PE for Pernambuco, and so on. The fatality rates were calculated from the daily number of deaths due to COVID-19, obtained from the health departments of each state through the official platform of the Ministry of Health [17]. All data is freezed at November 17th, 2021, and available in CSV format on Github [18] and the retrieve method is explained in the Supporting Information (Data Source). The total fatality up to this date was computed as 605,477. After the first few months of the pandemic, the numbers of deaths per State were large enough to render negligible their estimated statistical fluctuations due to the size of the samples. The voting rates are for the October 28th, 2018 election and are available on our Github page. This was the last round of the general election involving only the two most voted candidates in the first round. The total number of votes in the elected president was 57,797,847, of a total of 104,838,753 valid votes.

**Fig 2.**
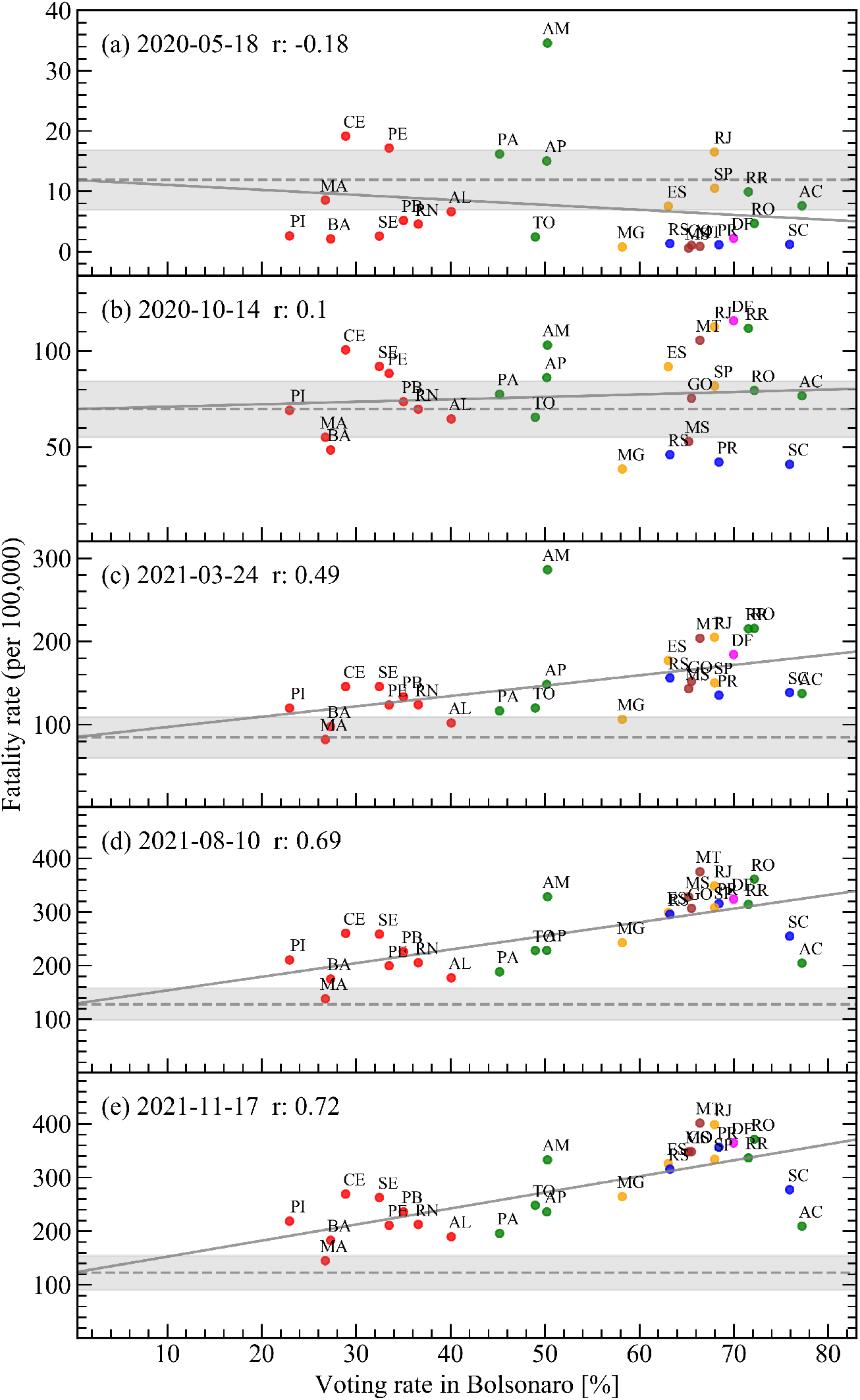
Distribution of fatality rates for all Brazilian States as a function of voting rate in the Brazilian president in the last election for five different days of the pandemic. The solid gray lines are linear fits to the data, and the dashed lines are the extrapolations to the fatality-rate level corresponding to a voting rate of 0%. The gray region is the error bar on the value of the dashed line. The colors for the State’s labels represent the same regions as in Fig. 1. *r* is the value of the Pearson correlation coefficient for the distributions in each panel.

At the beginning of the COVID-19 outbreak in Brazil (Fig. 2 panel a), there was no significant trend of the fatality rate with the voting rate in the Brazilian President. We can qualitatively verify this by adjusting a straight line to the points using the method of least squares, and obtaining both the inclination of the line and the point it crosses the vertical axis at 0%, with their respective error bars. The value the fit crosses the vertical axis is indicated by the dashed line, and its error bar is given by the gray area. Note that the straight line fitted to the distribution barely leaves the region of this error bar, indicating the low linear correlation of the distribution of points with the values on the horizontal axis. The observed small correlation was also negative. This initial negative trend can be understood from the regional correlations present in the distribution of voting rates itself. The smaller support for Bolsonaro came from the States on the Northeastern region (red dots in Fig. 2), the poorest, with smaller Human Developing Index (HDI), region of the country. The larger portion of vulnerable population and worse conditions of health care should result in larger death rates in these States once the pandemics hit the country, as observed in panel a of Fig. 2.

The correlation of voting rates in Bolsonaro with the richest, with larger HDI, States in the country is not related with Bolsonaro himself, but actually with his main adversary in the last election, the candidate from the Workers’ Party. As pointed by Bohn in Ref. [19], when the Workers’ Party started in the 1980’s, its electoral base was concentrated in the richer Southeastern region of Brazil, but this support slowly shifted to the regions in the country with lower HDI throughout the following decades. The correlation of support to the Workers’ Party with regions of smaller HDI is a consolidated trend observed since previous general elections in Brazil [20]. As the pandemics advanced in Brazil, we believe this trend of support to the opposition Party shielded, to a larger extent, the regions of the country with lower HDI from the political influence of Bolsonaro with respect to the pandemics.

As time passes by, then, the correlation in Fig. 2 becomes less negative and ends up turning positive around the date of panel b, inverting the initial trend dominated by the States HDI. The first traces of positive correlation with the vote in Bolsonaro started to be reported at the end of 2020 [21]. The correlation then continued to evolve to positive values, until it started to become significant around the date of panel c. First reports of its fast acceleration can be found then at the beginning of 2021 [22, 23]. Panels d and e finally depict the most recent situation of high correlation between the fatality rate in the Brazilian States and the respective voting rate in the Brazilian President.

These observations are quantitatively summarised in Fig. 3, where we plot the Pearson correlation coefficient *r* for the distributions of Fig. 2 for each day starting from the day of the first death, March 17th, 2020. Coefficients between −0.3 and +0.3 (blue region) indicate small or insignificant correlation [24]. Coefficients between +0.3 and +0.5 (green region) indicate a moderate positive correlation, and between +0.5 and +1.0 (red region) indicate a strong positive correlation. Figure 3 depicts, then, the onset over time of a strong correlation between fatality rate and the political preferences of the various Brazilian States, as inferred from the results of the country’s last general election. The establishment of such strong correlation coincides with the period over time, between the two vertical dashed lines, where the Health Minister for the central government in Brazil, General Eduardo Pazuello, was completely aligned with the views of the Brazilian President on how to deal with the COVID-19 crisis. This was not the case in the very beginning of the pandemic in Brazil, when the actions of the Presidency against the measures to control the pandemic were often conducted against the counseling of the Health Ministry, which led twice to the substitution of the head of the Ministry, up to the nomination of General Pazuello in May 2020. As an example of the significance of this timeline, five days after assuming the post of acting Health Minister, his Ministry released an informative note [25] with orientations on the use of a series of medications whose efficacy have not been previously established for the treatment of COVID-19. This untested treatment was explicitly advertised by the President and received increased support of the Ministry of Health, as tested-and-proved measures like mask mandates and social distancing were systematically downplayed [4].

**Fig 3.**
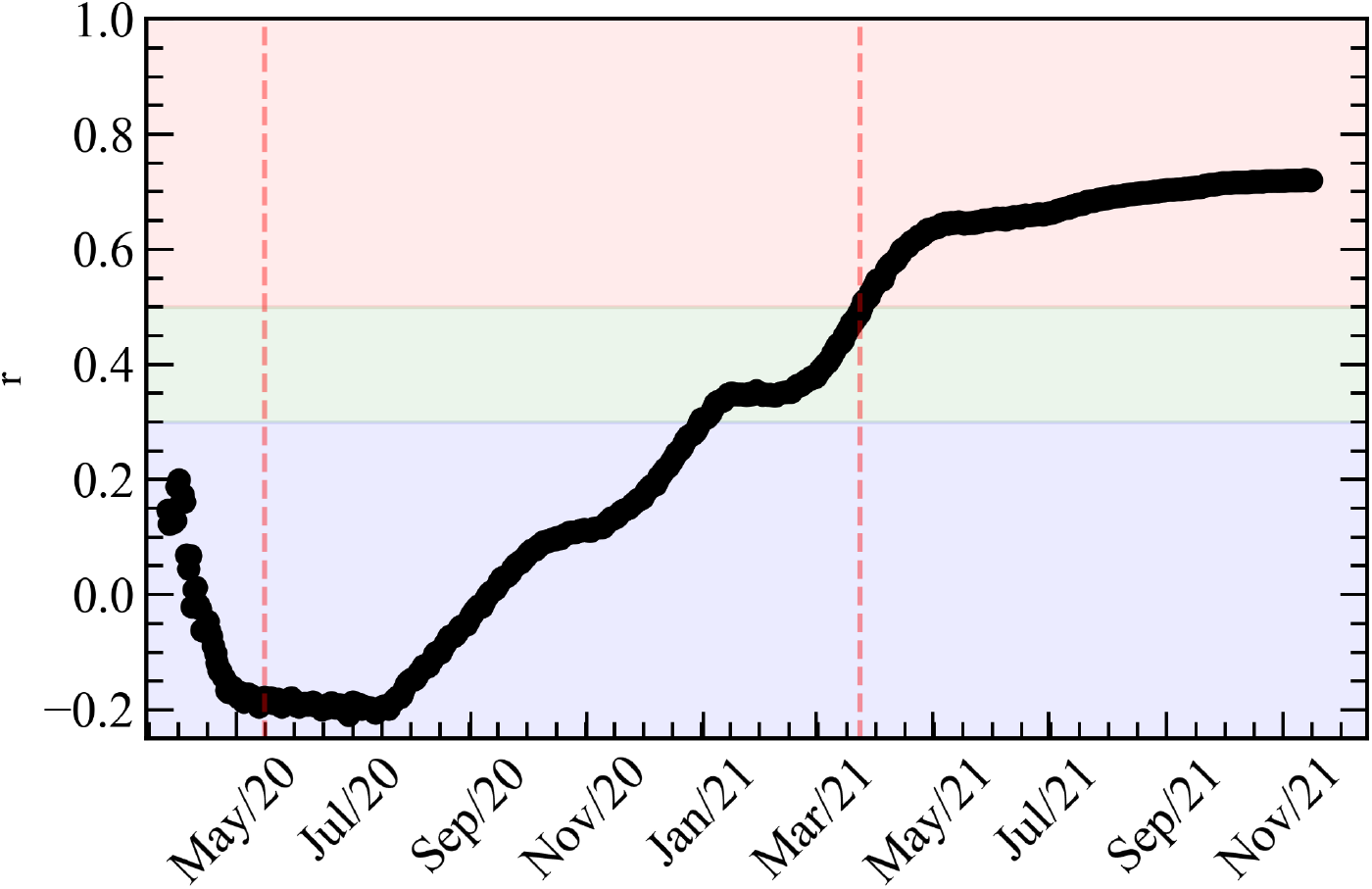
Evolution of correlation in Brazil between the States fatality rates and its voting rate in the Brazilian President, as measured by Pearson’s *r* coefficient. The vertical dashed lines indicate the beginning and ending of the period in which General Eudardo Pazuello was head of the Health Ministry.

## 2 Results

The correlation coefficient in Fig. 3 (see Supporting Information Pearson correlation coefficient) demonstrates the strong influence the political narrative had in the death rates as the pandemic developed in Brazil. This correlation coefficient may be translated into an excess of total fatality in the country due to political influence, a more concrete measure of the overall impact this process had in Brazil so far. Excess of fatality due to any particular cause is defined as the difference between the observed fatality and an estimate of fatality without such cause. In a health crisis, this estimate is commonly done by comparing the fatality during the crisis with that of previous years. In our case, however, we need a different approach, since we need to estimate what the fatality during the COVID-19 crisis in Brazil would be without the political factor, and then subtract this number from the observed fatality to obtain the excess fatality. From the above discussion, however, this estimation can actually be done straightforwardly by computing first the total fatality coming from the level of the fatality rates indicated by the dashed lines in Fig. 2, i.e., the level corresponding to 0% of votes in Bolsonaro. This provides an estimation for the expected number of deaths if there was no correlation with the voting rate in the Brazilian President. This estimation is then subtracted from the total number of COVID-19 death, and the result is our estimation for the excess in total fatality in Brazil due to political influence. This number as a function of time is then plotted in Fig. 4. The error bars in the figure come from the gray regions in Fig. 2.

**Fig 4.**
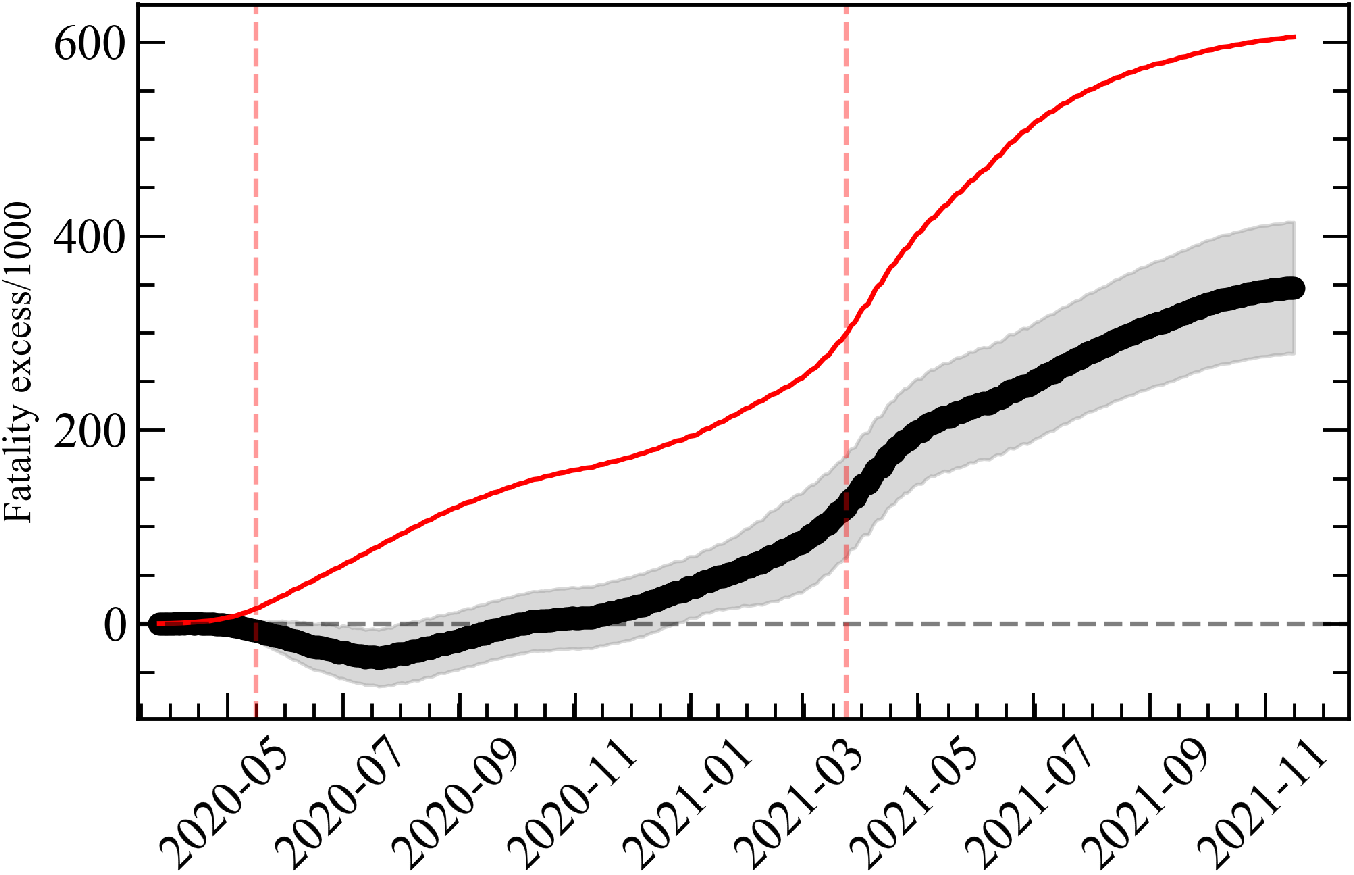
Evolution in time of the total number of deaths in Brazil due to COVID-19 (red line) and of the excess of fatality in Brazil due to the political influence of its President (black line). Error bars of the black curve are given by the gray region. The vertical dashed lines indicate the beginning and ending of the period in which General Eduardo Pazuello was head of the Health Ministry.

From Fig. 4 we note that the growing correlation only starts to translate into a sensible excess fatality around December, 2020. From that point onward, however, the political correlation starts to dominate the total fatalities due to COVID-19 in Brazil, until it reaches a total excess fatality of 350 ± 70 thousands on 17 November, 2021. This means that we estimate that (57 ± 11)% of deaths attributed to COVID-19 in Brazil are related to the political influence the President had over the Brazilian population in the period of the pandemic. Another conclusion we can draw from the observed dynamics of the pandemic in Brazil is that this political influence ended up surpassing, in the last months, all other possible causes of its aggravation in the country, like the new strains of the virus. Finally, as it dominated its last stage, this wave of politically-influenced deaths postponed for months the containment of the pandemic in Brazil, enhancing its other socioeconomic impacts.

The above estimation, however, has still its drawbacks. First, our estimation calculates the excess deaths compared to a zero-correlation level, while Fig. 3 shows that we should actually expect a negative correlation coming from the poverty level of the States that least supported the President. This effect is responsible for the negative part in the beginning of the excess-of-deaths curve in Fig. 4. On the other hand, this negative dip can also serve as an estimation for how much we are underestimating the number of deaths by assuming zero correlation as our baseline. From the minimum value of this dip, we expect that corrections due to a negative-correlation baseline would be smaller or on the order of our final error bar. A second effect we cannot estimate is any excess of deaths resulting from delays in the beginning of the vaccination program by the Federal Government, since this affected all States equally. Finally, the excess of deaths due to the a lack of a coordinated national response to the crisis was estimated in Ref. [14] to be around 120 thousands deaths. This number is not completely independent of our estimation, since there was some degree of coordination among the Northeastern States in their approach to the pandemics, whose positive effect would reinforce the final correlations present in the above figures. This regional effort, however, was largely independent of the rest of the country and could not completely compensate the lack of a national strategy.

## 3 Conclusions

The scientific method, in a nutshell, applies reason and common sense to search for the best response to specific problems. In the case of the ongoing pandemic, the presented problem was how to minimize the deaths related to the spread of COVID-19. In this way, since the beginning of 2020, many new treatments were tested, specific sanitary and social distancing protocols were established, and various effective vaccines were developed in record time. In this context, as a significant portion of the political forces in Brazil chose to ignore such fast-paced development and settled with a mixture of wishful thinking and untested treatments, the consequences were dire, directly resulting in an excess of deaths among populations sharing these political views.

Our analysis, in summary, shed a new light on the role that broad political views may have in severe health crisis. It reveals, specifically, the somewhat unexpected magnitude of such political bias over the spread and fatality of the pandemic in Brazil, overcoming at a certain point in time other strong factors such as poverty levels and the mutation dynamics of the virus itself. As the vaccinated population grows in Brazil and the pandemic seems to finally lose its strength, we hope the Brazilian case in the COVID-19 pandemic will serve as a warning in future health and environmental crises as to the dangers of ignoring informed, sensible advices formulated through the fair application of the scientific method.

## Data Availability

All data and code used can be found at https://github.com/monolipo/politicalcovid19

## Supporting information

### Data Source

The data was retrieved from TSE using the website “Gazeta do Povo” and compiled at: https://especiais.gazetadopovo.com.br/eleicoes/2018/resultados/mapa-eleitoral-de-presidente-por-municipios-2turno

### COVID19 Deaths confirmed by state

All cases reported here were confirmed by the health departments of each state, and also obtained by the official platform of the Ministry of Health. All data is available in CSV format on Github: https://github.com/wcota/covid19br [17]. The fatality data and votes were related using the Pearson Coefficient and the code is avaliable at [18].

### Pearson correlation coefficient

The Pearson correlation coefficient is named for Karl Pearson and gives us the strength of the linear relationship between two data samples. It is calculated as the covariance of the two variables divided by the product of the standard deviation of each data sample. It is the normalization of the covariance between the two variables to give an interpretable score:

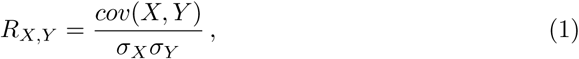

where *cov*(*X, Y*) is the covariance between X and Y,

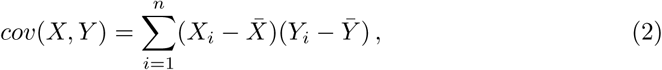

with *n* for the sample’s size, *X*_*i*_ and *Y*_*i*_ the individual sample points and 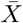 and 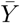 the mean of samples X and Y. As the standard variation is simply the variance squared, the Pearson Correlation Coefficient is then:

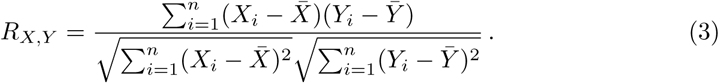

The coefficient returns a value between −1 and 1 that represents the limits of correlation from a full negative correlation to a full positive correlation. A value of 0 means no correlation. The value must be interpreted, where often a value below −0.5 or above 0.5 indicates a notable correlation, and values below those values suggests a less notable correlation.

To get the Pearson correlation coefficient for our data, we tested the SciPy function scipy.stats.pearsonr, wich gives us the correct value for each date, and also the np.corrcoef from numpy wich gives the same result.

## Notes

### Competing Interest Statement

The authors have declared no competing interest.

### Funding Statement

The authors received no specific funding for this work.

### Summary of Updates

Affiliations corrected

